# Using wearable and nearable devices in telerehabilitation for COPD: A review of digital endpoints in home-based programs

**DOI:** 10.1101/2025.09.02.25334970

**Authors:** Stephanie Zawada, Louis Faust, Caden Collins, Moein Enayati, Roberto Benzo, Emma Fortune

## Abstract

**Objective:** Despite its demonstrated effectiveness at improving outcomes, pulmonary rehabilitation (PR) for chronic obstructive pulmonary disease (COPD) is underutilized. Sensor-generated data from wearable devices have the potential to mitigate this challenge by generating digital endpoints that provide insights into patient behaviors at home; however, there is no consensus on how to measure home-based PR (HBPR) outcomes with these tools. This review aims to describe (1) the most frequent digital endpoints used in HBPR studies and (2) the devices used to capture these endpoints, summarizing gaps in their applications to HBPR for COPD patients.

**Methods:** We completed a scoping review using the PRISMA checklist across databases (Web of Science, Scopus, and OVID) from January 2005 to June 20, 2025. We included peer-reviewed articles on HBPR for COPD, excluding reviews, commentaries/editorials, poster abstracts, and conference proceedings. Eligible articles included cohort studies and clinical trials of adult patients (age ≥ 18 years) with COPD participating in HBPR that include one or more digital endpoints.

**Results:** Among eligible articles (n = 218), 13 (6.0%) met inclusion criteria, the majority of which were published after 2020 (61.5%). Most studies enrolled fewer than 100 COPD patients (76.9%) for an average monitoring period of 12.5 weeks. Activity trackers were the most commonly used device (46.2%) to capture data. The most frequently used digital endpoints were step count (84.6%), time spent active (38.5%), and time spent sedentary (30.8%). Two study designs were used: randomized controlled trial (76.9%) and observational cohort. Study designs were heterogenous with more than one-third (38.5%) presenting a lack of statistically significant results.

**Conclusions:** Although we identified analogous digital endpoints in some studies, dissimilar methods and study designs remain barriers to synthesizing results generated from HBPR programs for COPD. Wearable devices have the potential to build novel PR models, but more work is needed to translate real-world data into clinically meaningful measures. Future research should elucidate which participants would benefit most from and complete HBPR to build an evidence base for the validation of HBPR-relevant digital endpoints, particularly those derived from less common sources like cardiovascular and sleep measures.

**Clinical Implications:** While wearable devices can augment HBPR programs, caution must be exercised when interpreting remotely collected data and requires clinicians to engage patients via traditional monitoring means, such as video telehealth and in-person visits, to ensure accurate screening and severity grading. Combining multiple sensors can capture a range of physiologic markers relevant to understanding patient-specific COPD exacerbations, from medication use to sedentary behavior, potentially informing personalized rehabilitation plans. Considering the heterogeneity of randomized controlled trials with digital endpoints in HBPR programs, little is known about optimal COPD patient sampling frequencies, or which populations would benefit most from integrating specific sensors in home-based programs.

## INTRODUCTION

Chronic obstructive pulmonary disease (COPD) is a leading cause of death worldwide, and its prevalence is projected to increase by 2030 [1,2]. Exacerbations of COPD, characterized by worsening of chronic breathing problems, are a major cause of hospitalization and death. In addition, these episodes place significant stress on the heart, contributing to the estimated 60% of COPD-related deaths attributed to cardiovascular disease [3]. Considering that 65% of patients with COPD are overweight or obese, the need for scalable, effective prevention strategies to lower cardiopulmonary risk factors, like sedentary behavior, in this population is urgent [3]. Pulmonary rehabilitation (PR) is one such approach to increase physical activity, reduce shortness of breath, and enhance mood in patients with COPD [4,5].

PR is a well-established treatment, effective for improving outcomes and quality-of-life (QoL) after hospitalization for COPD exacerbations [6]. Though PR for COPD can be personalized to individual patient needs, central elements of PR involve endurance training, strength-building exercises, and breathing techniques [7,8]. These components are coordinated by clinical teams, including nurses, respiratory therapists, physical therapists, and counselors, and, sometimes, by autonomous tools [7]. PR for COPD typically consists of three phases, each with ongoing education about how to manage COPD. Phase I (inpatient) occurs while a patient receives care for a COPD-related hospitalization. Phase II (outpatient) is delivered multiple times per week for 6 or more weeks with clinical team oversight. Phase III (maintenance) is an extension of Phase II with less clinical oversight, as patients transition from monitored exercise and treatment to self-management [9].

Enrolling COPD patients in outpatient PR is supported by current evidence, including the majority of observational studies, randomized controlled trials (RCTs), and systematic reviews [10–14]. Despite its positive effects, outpatient PR programs remain underutilized, with recent studies showing fewer than 4% of patients hospitalized for COPD participating in PR after discharge [15,16]. Persistent reasons cited by patients who decline PR are a lack of motivation, interest, and perceived benefit [17–19]. The transportation burden associated with PR also drives this lack of adoption, with in-person PR primarily concentrated in urban environments [20]. Thus, access disparities are worse in rural areas, with fewer than 12% of rural patients having a PR center within 10 miles from their home [21]. Limited insurance reimbursement for PR is an additional challenge for PR programs, exacerbating the already-low participation rates observed in low socioeconomic status (SES) patients [22–24]. For those who do enroll in PR, attrition rates are high, often exceeding 50% [25].

To address these barriers, alternative, home-based-PR (HBPR) models that rely on telehealth technologies, have been implemented [26], however, no specific model of HBPR has been defined, with the clinical practice guidelines (2023) from the American Thoracic Society (ATS) on PR in COPD stating only that patients with stable COPD should be offered center- or home-based PR options [27]. Similarly, a 2021 Cochrane review, including 5 randomized controlled trials (RCTs) and 2 controlled clinical trials (CCTs), compared center-based PR with HBPR, the latter limited to only telephone and videoconferencing modalities [28]. Their results found clinically meaningful improvements in exercise capacity, QoL, and dyspnea in both groups and no significant differences in improvements between groups. Notably, participants in HBPR programs were more likely to complete PR than those in center-based programs. Many of these programs offer flexibility beyond that of in-person PR, possibly addressing lifestyle needs and behavior changes critical to improving adherence [29]. Moreover, when these programs collect real-time data from wearable devices, they offer clinical teams the opportunity to tailor patient-specific exercise prescriptions and behavioral interventions during outpatient as well as self-management PR phases for sustained impact [6,30–32].

Several systematic reviews have assessed the effectiveness of wearables for monitoring COPD patients, with none specific to outpatient PR programs. A 2023 meta-analysis (n = 37) by Shah et al. showed that wearables, consisting of mostly pedometers, increased mean daily step count and performance on the 6-minute walk distance test over short time periods [31]. Moreover, wearables combined with PR programs showed improvements of greater magnitude compared to remote monitoring without coaching or observation, corroborating the results of a 2016 meta-analysis assessing older technologies [33]. The use of wearables data to predict or avoid respiratory exacerbations in COPD is indeterminate and had no significant impact on QoL, extending the results of a systematic review of predictive algorithms for at-home COPD monitoring [34]. Additionally, a systematic review (included papers, n = 7) assessing oxygen saturation and respiratory rate readings from Bluetooth-compatible devices reported high validity in real-world settings, but yielded inconsistent prediction accuracy [35]. Additionally, numerous proof-of-concept studies for wearables have only been conducted in controlled environments, like inpatient PR, not accounting for the complexities inherent with real-world deployment [36–37]. No review exclusively focused on HBPR settings has assessed wearable devices. Moreover, the reviews available assess the outpatient and self-management PR phases together, without considering that COPD patients in outpatient PR are normally in a higher-acuity state than those in the self-management PR phase [38,31].

Despite the promise of objective data from wearable devices, further work is necessary to translate these measures into clinical practice. To our knowledge, no review has surveyed the wearables landscape in HBPR to map the state of currently available digital endpoints: measurable health outcomes from digital technologies used to evaluate whether an intervention has a targeted effect [39]. This scoping review primarily aims to identify the set of digital endpoints used to study patient outcomes in HBPR programs for COPD. Second, it aims to summarize gaps in our understanding of using wearable and nearable devices in HBPR to monitor COPD.

## METHODS

### Definitions and analytic framework

To organize this scoping review, we conceptualized the HBPR program as monitoring or delivering an intervention to patients with a COPD diagnosis, facilitated by digital sensors embedded in wearable or nearable devices. Included studies were those investigating fully remote outpatient PR implementations. Studies with home health visits and in-person assessments, except in a preliminary phase before HBPR or to provide orientation for HBPR, were excluded. Studies with telehealth visits were eligible for inclusion, provided some digital sensor from a wearable sensor device was a monitoring component of the program.

Eligible studies were those assessing at least one digital endpoint, like steps per day. Digital endpoints were derived from sensors in a wearable or nearable device and could capture any physical symptom capable of being observed remotely. No specific definitions for devices were applied to limit the scope of emerging technologies included in this analysis. For instance, smartphone app and integrated smartphone app-platform technologies were included if they collected sensor-based data, like smartphone accelerometer data.

We applied this framework to categorize types of digital endpoints and summarize their state-of-the-art implementations in real-world PR programs with COPD patients, thereby highlighting gaps in the literature. Although prior frameworks informed by theory have been proposed to evaluate state-of-the-art technologies for clinical applications, few have focused on assessing digital endpoints. Our approach builds on that of Goldsack et al.’s framework for digital endpoint evaluation. We assembled the set of digital endpoints able to be observed in real-world settings and the devices used to capture them [40,41].

### Search Strategy

This review was conducted according to the PRISMA Extension for Scoping Review guidelines. Searches were conducted across Web of Science core collection (hosted by Clarivate Analytics [≥1975]), Scopus (hosted by Elsevier [≥1788]), and Ovid MEDLINE (≥1946), in addition to Epub ahead of print, in-process, and other nonindexed citations and daily, which is identical to PubMed. Studies published from January 2005 through June 20, 2025 were included. Only publications available in English were included. Observational studies, including both cohort and case-control studies, as well as cross-sectional and mixed-methods studies were eligible for inclusion. Randomized (RCTs) and clinical controlled trials (CCTs) were eligible for inclusion.

Reviews, conference abstracts and papers, commentaries/editorials, protocols, and professional society guidelines were excluded. As the scope of this review centered on real-world data capture of symptoms and markers relevant to outpatient PR for COPD, prototype studies as well as those assessing only PR program adherence and usability metrics were excluded. Eligible studies included adults (≥ 18 years of age) with a COPD diagnosis.

Limiting our search strategy to only studies with “chronic obstructive pulmonary disease” or “COPD” in the title or abstract, a robust set of digital sensor tools were included in the search terms (Figure 1).

**Figure 1.**
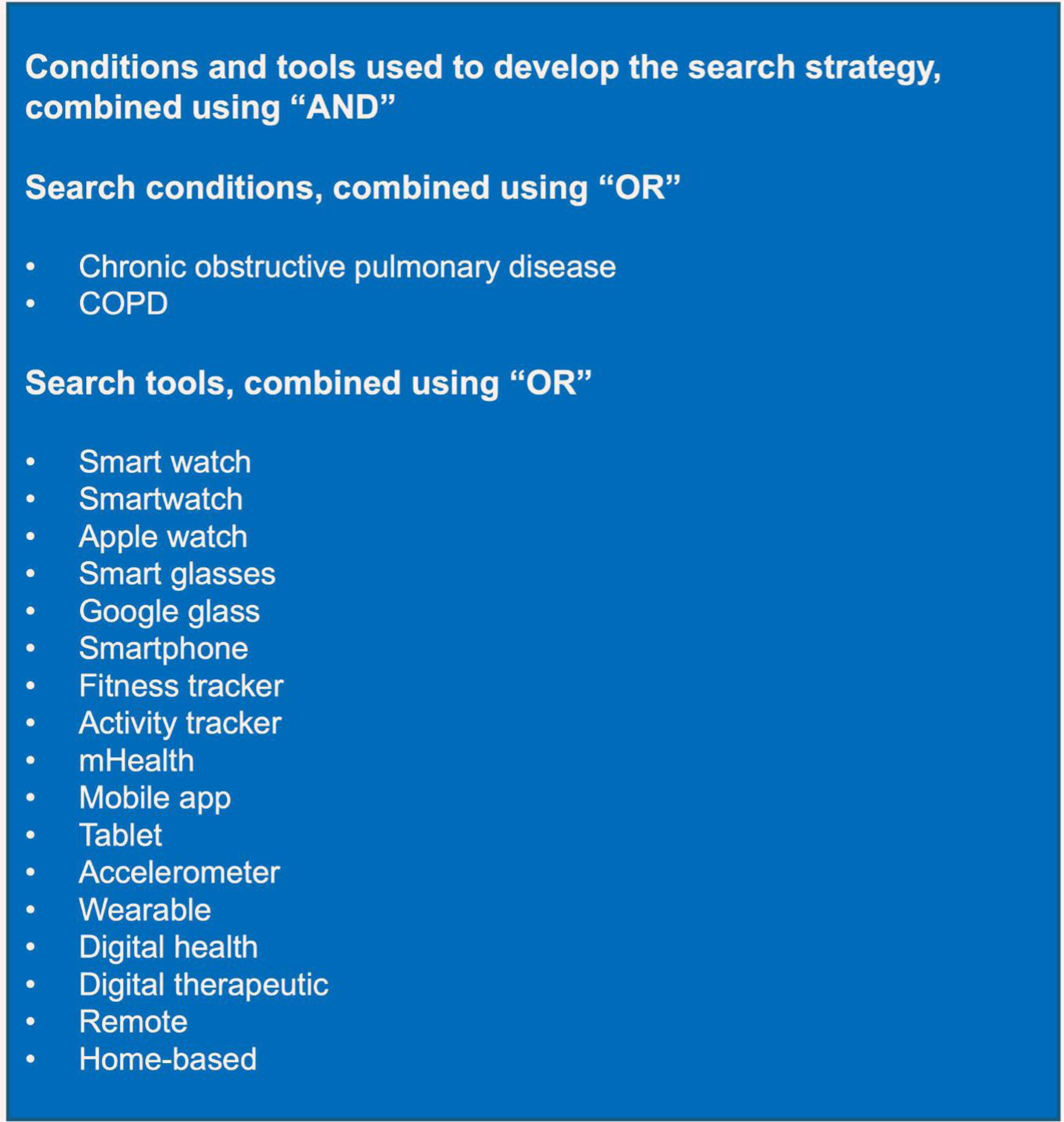
Conditions and tools used to develop the search strategy.

### Selecting Studies

First, title and abstract screening for relevance was performed by one investigator (SJZ). At this step, the list of studies was reviewed by 3 additional investigators (JVC, LJF, and EF). Second, a full-text review, including annotation and adjudication, was completed by two investigators (SJZ and CAC). The final list of included studies was reviewed by a clinical expert to ensure relevance (RJB). The rationale for each eligibility criteria applied is available in Table 1.

**Table 1.** Eligibility Criteria and Rationale for Study Inclusion and Exclusion

| Eligibility criteria and variable | Rationale |
| --- | --- |
|  | <u>Inclusion criteria</u> |
| Outpatient PR phase | Participants must be participating in an outpatient PR program, with data collection at the beginning, end, or during the program |
| Fully remote setting | Data must be collected outside of clinical environments and in real-world settings |
| Participants exclusively with a COPD diagnosis in the observational or experimental cohort | Participants must be diagnosed with COPD, as evidenced by clinical documentation or assessment |
| Peer-reviewed research | Only studies published in peer-review journals will be included, due to their credibility |
| Wearable or nearable device | Tools used to collect digital endpoint data must be from a wearable device, like a smartwatch, or a nearable device, like a metered inhaler |
| Empirical study design with observational, comparative, and/or clinical trial components | Studies must address a hypothesis or research question |
| Published between January 2005 and June 20, 2025 | Included studies conducted over the past two decades to provide a comprehensive analysis of all digital endpoints proposed |
| Articles published in English | Limited due to language proficiency of investigators |
| Participant sample data included | Articles must include the number of participants, at minimum |
| **<u>Exclusion criteria</u>** |  |
| Studies including participants without COPD diagnosis | Aside from healthy controls, studies must include participants with COPD diagnoses to minimize disease-related biases |
| Nonempirical studies | Editorials, guidelines, reviews, brief reports, and other articles without a clear hypothesis or research question were excluded to minimize viewpoint bias |
| Studies focused exclusively on feasibility, usability, acceptability, or adherence | Though pilot and feasibility studies with explicit research questions involving digital endpoints were eligible for inclusion, studies exclusively focused on proof-of-concept or qualitative work were excluded from this review of sensor-generated digital endpoints |
| Studies focused on inpatient (Phase 1) or self-management (Phase 3) phases of PR | Studies focusing on phase 1 or 3 of PR were excluded |
| Device or algorithm validation studies | Considering this review aimed to map the state-of-the-art in digital endpoints for remote PR in COPD patients, studies proposing new devices or predictive algorithms were not appropriate for inclusion |
| Wearable or nearable device implementation with no sensor-based data analyzed | Given that this review focused on digital endpoints, any studies where wearable and nearable devices were used during PR but their resulting data not analyzed were excluded |
| Studies focused exclusively on qualitative data, including patient-reported surveys submitted via nearable and wearable devices | Studies with only qualitative data or only patient-reported surveys, such as those submitted via smartphone apps, were excluded, as they are not sensor-based endpoints |

**Table 2.**
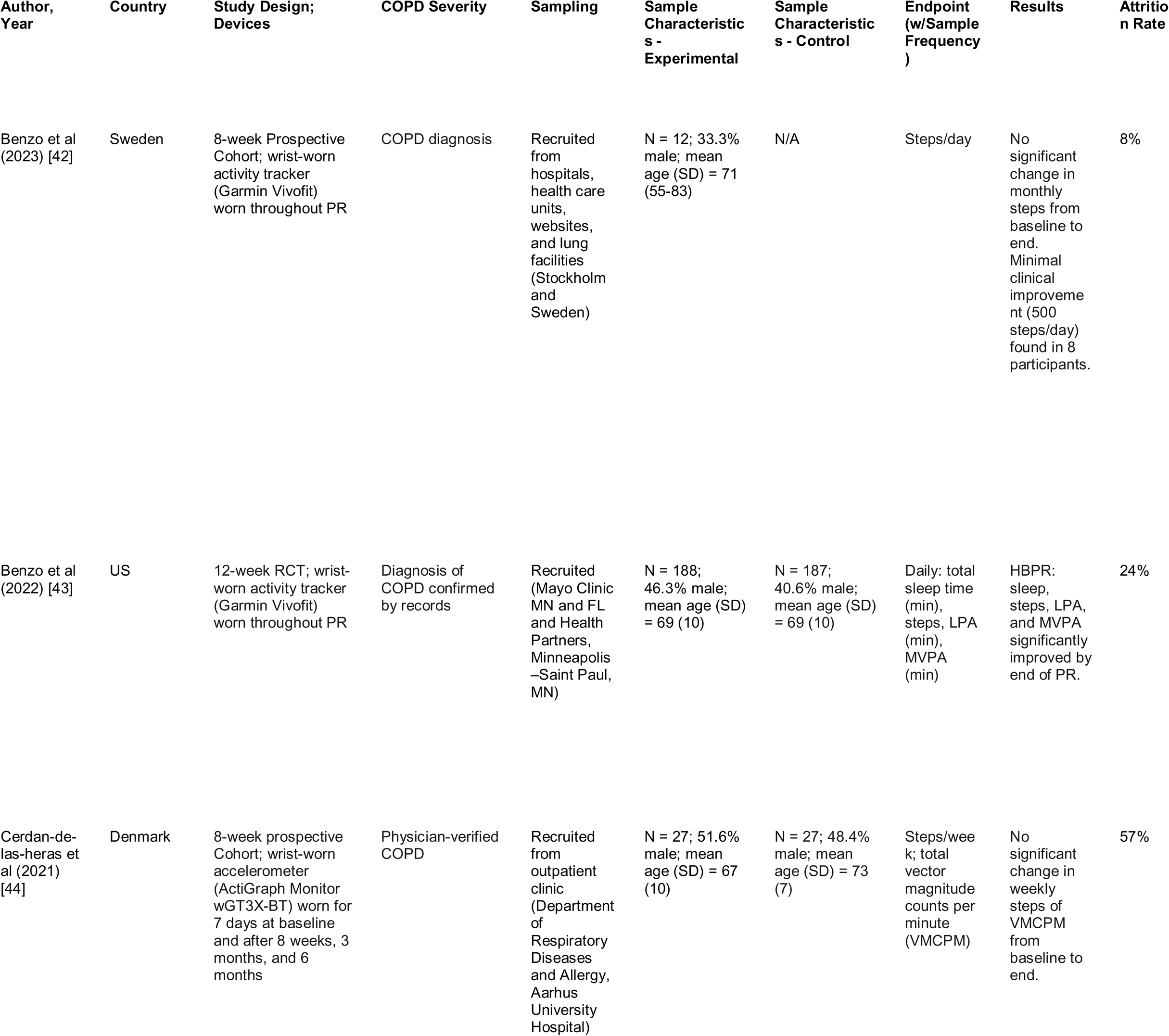
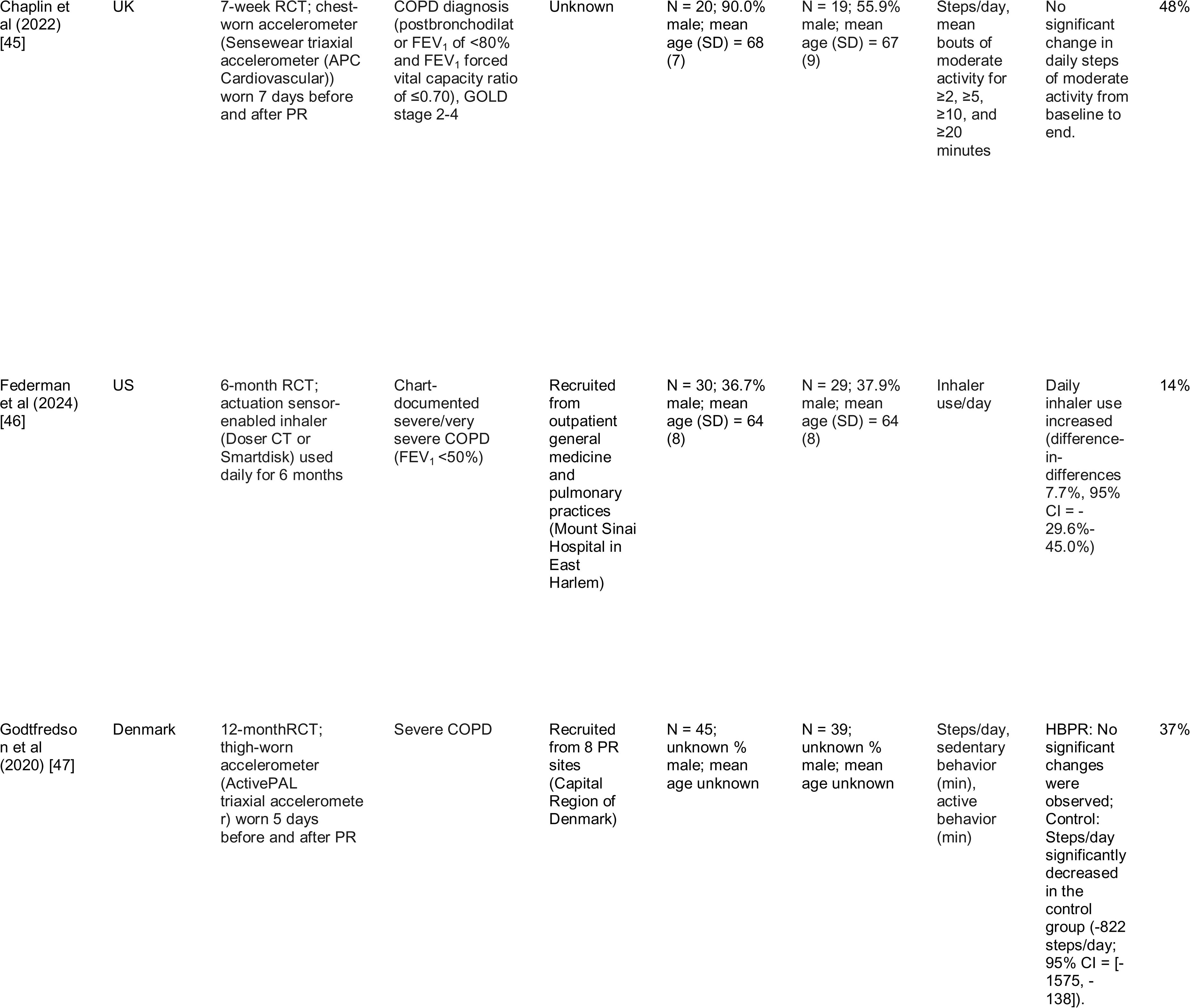
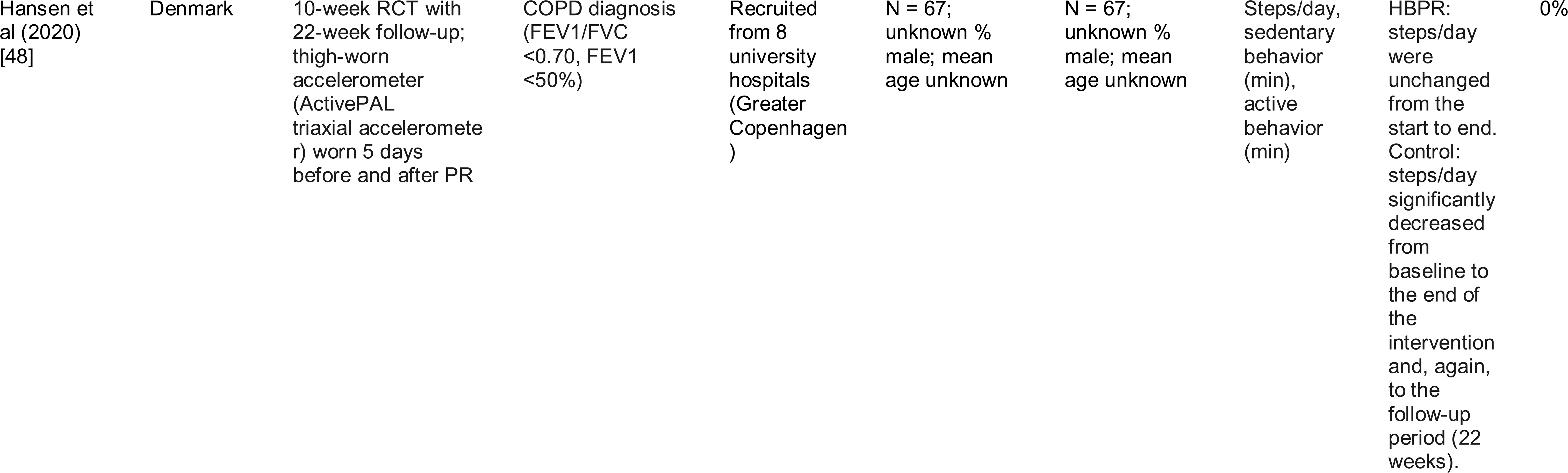
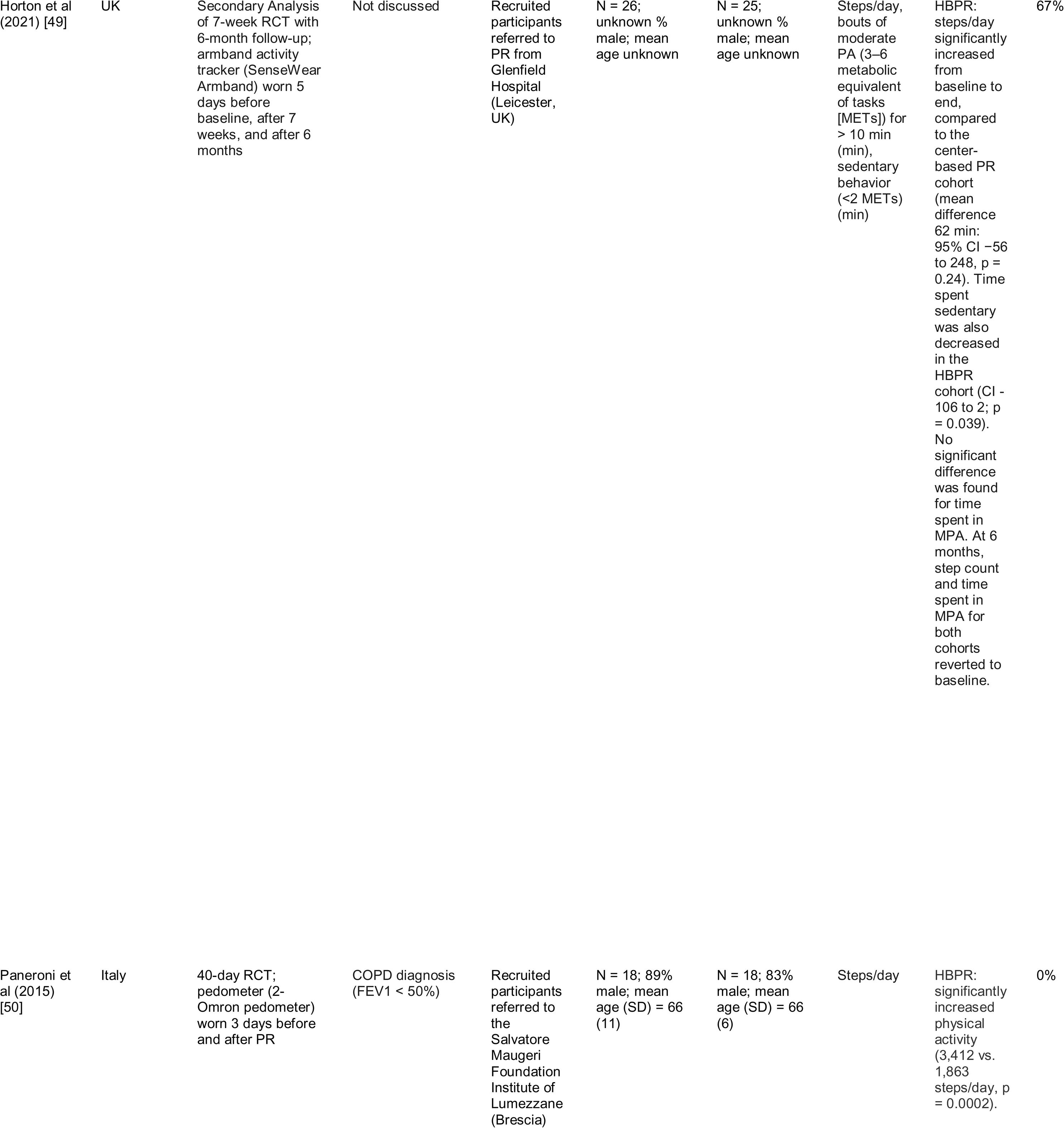
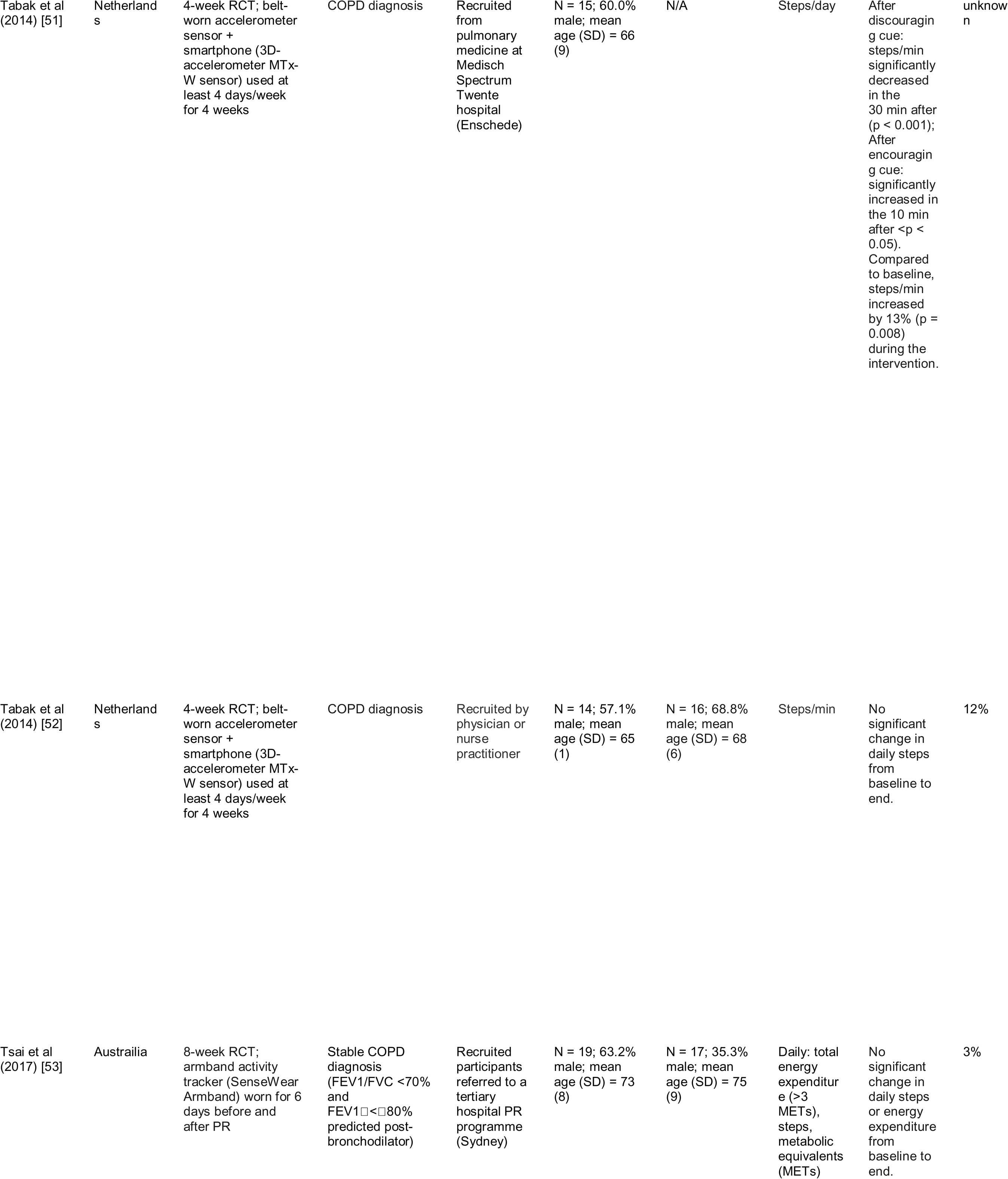
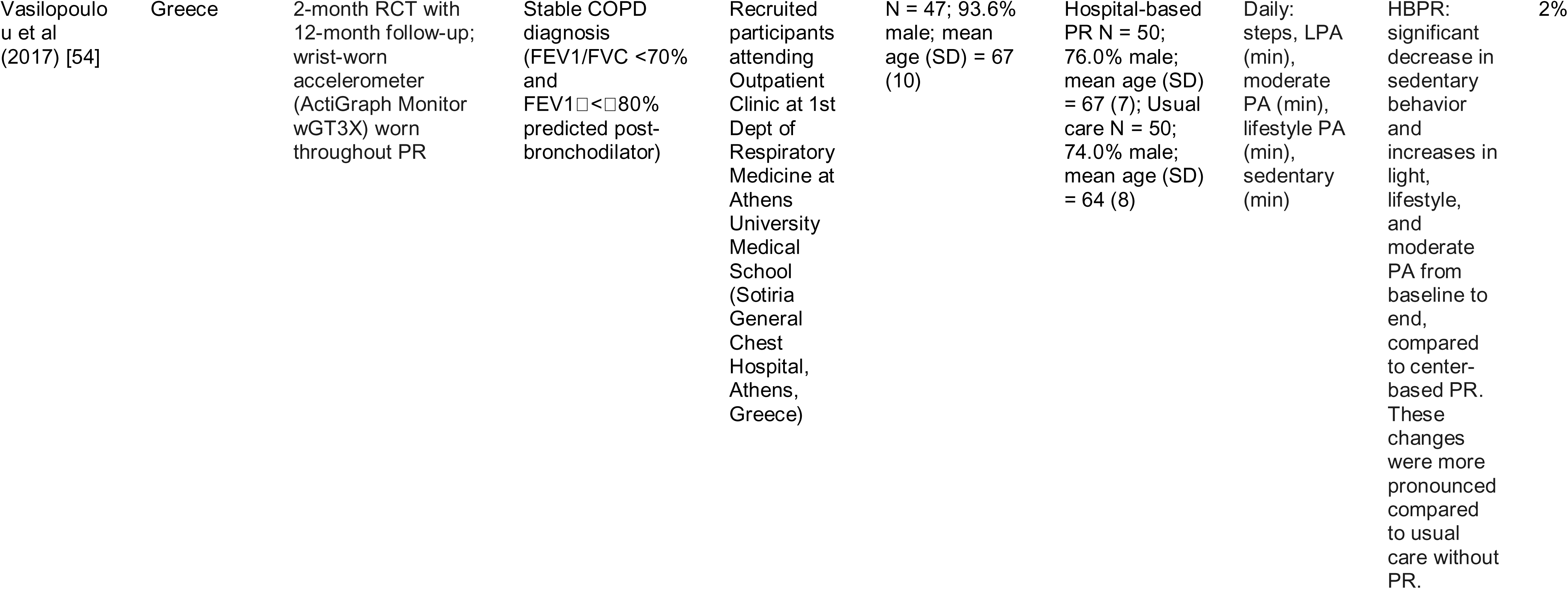
Summary of Studies on Digital Endpoints in HBPR for COPD

### Data Collection and Analysis

The following data was extracted by one investigator (SJZ): authors, year, country, study design, COPD severity, sample characteristics, wearable/nearable device, digital endpoints, results, and attrition rate. When reported in studies, non-digital endpoints, like patient-generated surveys or in-person walking tests, were not extracted for evaluation. Similarly, when a wearable or nearable device was used in a study but no recorded data were analyzed, the study was excluded. Digital measures used in descriptive studies were excluded from this analysis of digital endpoints. Smartphones were only listed as a monitoring device used to generate digital endpoints if they enabled the collection of sensor-based data, regardless of the smartphone’s role in the study.

## RESULTS

This initial search generated 14,895 articles. After removing duplicate articles, and then screening titles and abstracts, 218 titles were selected for full-text review. Of the 218 articles adjudicated, 6.0% (n = 13) met the inclusion criteria. The PRISMA flow chart for this search is detailed in Figure 2.

**Figure 2.**
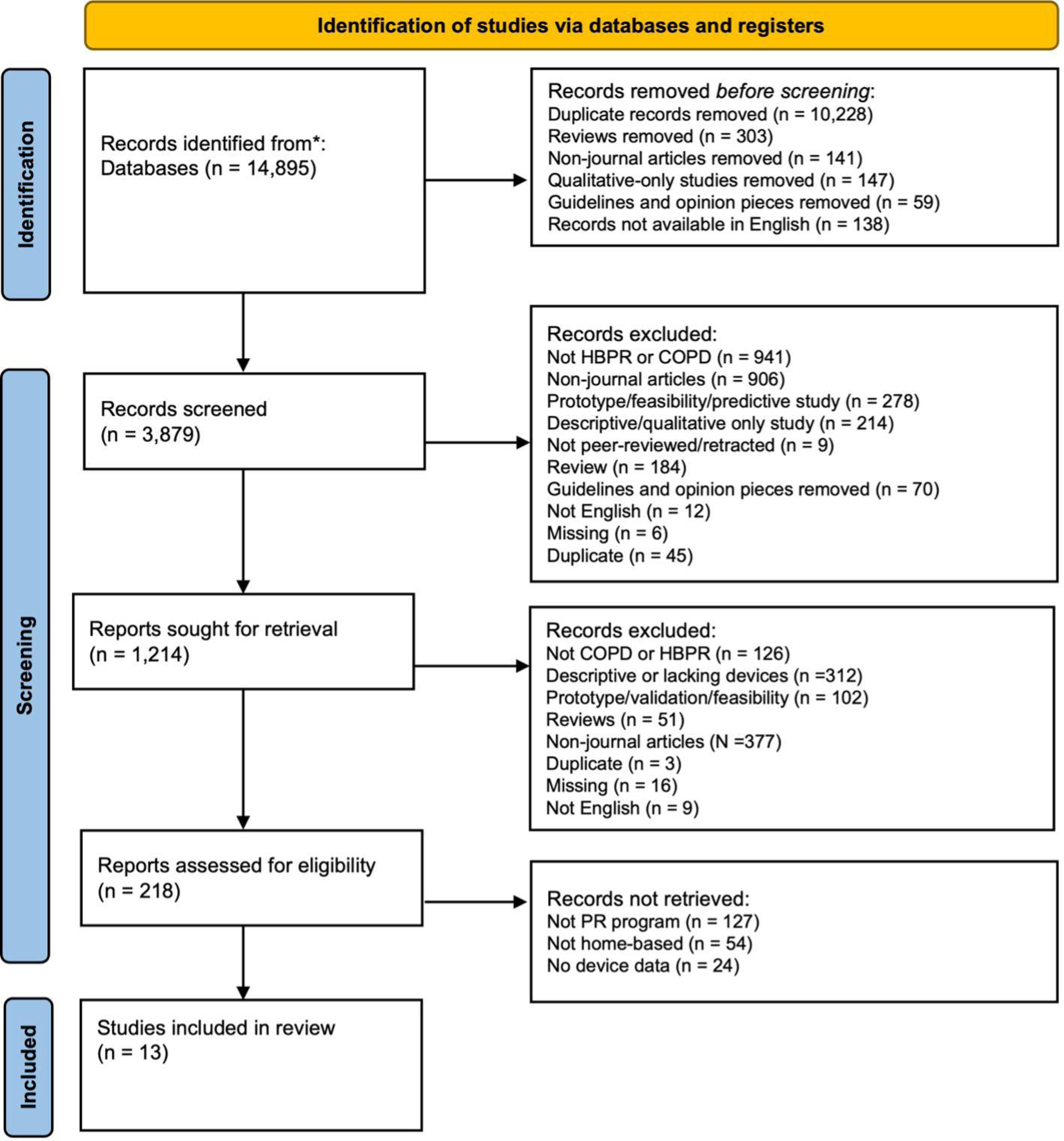
PRISMA Flowchart of Included Studies

### Study Characteristics

Summary data for each study is reported in Table 1. Most studies (n = 8, 61.5%) were published after 2019. For study design, 10 articles reported a randomized controlled trial (RCT), 2 used an observational cohort, and 1 was a secondary analysis of an RCT. More than half of the studies (69.2%) enrolled small cohorts of COPD patients (<100). The average duration of PR for studies was 12.5 weeks (min = 4 weeks; max = 52 weeks). One study had a 6-month follow-up checkpoint and another had a 12-month checkpoint. All studies focused on adult populations, primarily consisting of middle-aged and older adults, with the youngest cohort mean age of 64 years. Three studies lacked demographic enrollment data [47–49]. Of the studies with participant demographic data, half enrolled primarily male participants.

### What are the most frequently used digital endpoints in HBPR for COPD?

Among the articles included, all endpoints were generated using daily summaries of wearable data. Five studies (38.5%) examined more than 1 digital endpoint listed in Figure 3 [43,46–48,53,54]. Step count was the most common endpoint examined (84.6%), with most studies assessing steps/day and only 1 tracking steps/min. The next most popular endpoint was minutes spent in active behavior (38.5%), minutes spent in sedentary behavior (30.8%), minutes spent in moderate physical activity (23.1%), minutes spent in light physical activity (15.4%), minutes spent in total sleep (7.7%), minutes spent in moderate-to-vigorous physical activity (MVPA) (7.7%), minutes spent in lifestyle behavior (7.7%), and inhaler use (7.7%). One implementation of the “minutes spent in active behavior” endpoint was assessed within the 30 minutes of a motivational cue (intervention) delivered to patients via smartphone [52].

**Figure 3.**
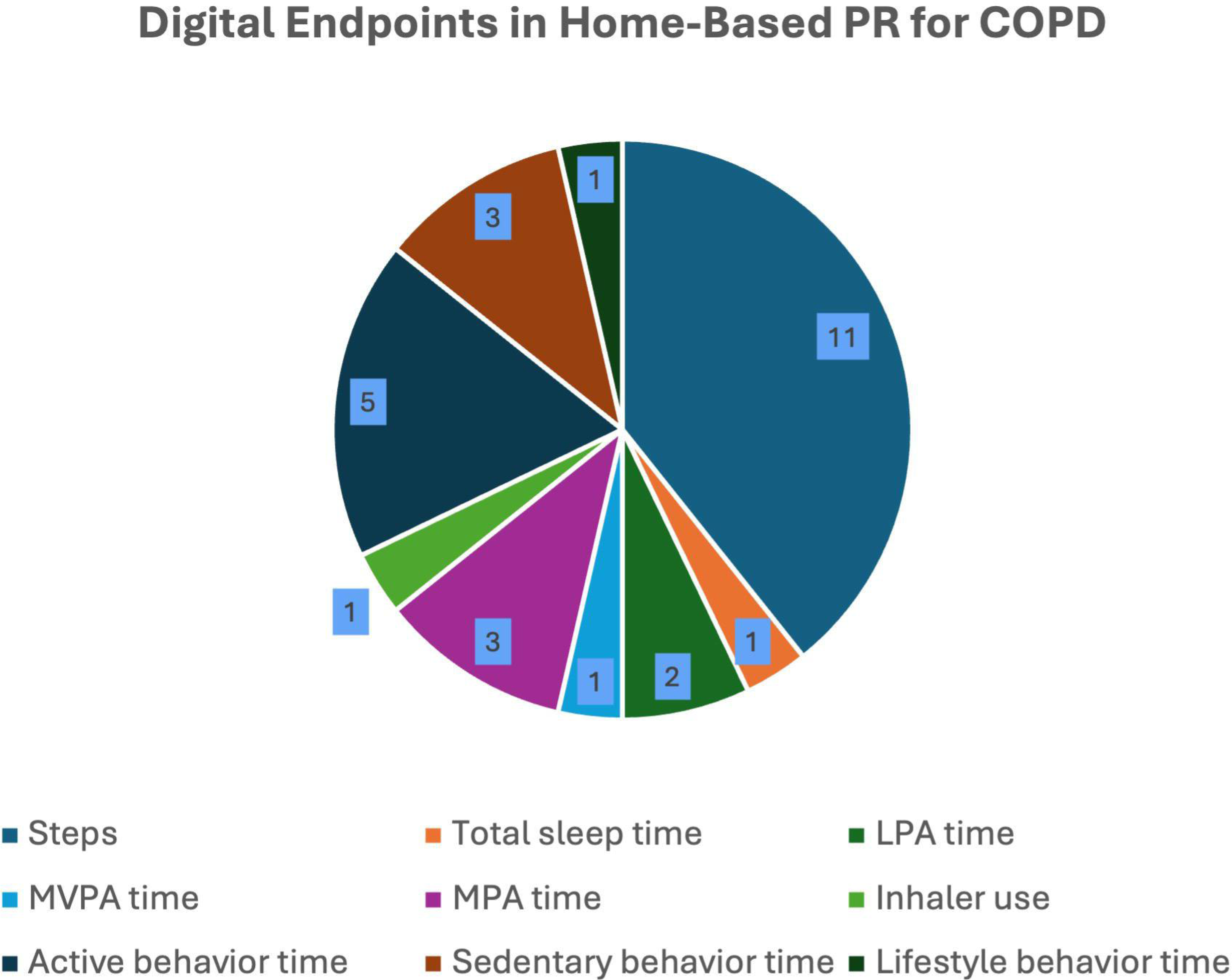
Frequency of Digital Endpoints Used

**Figure 4.**
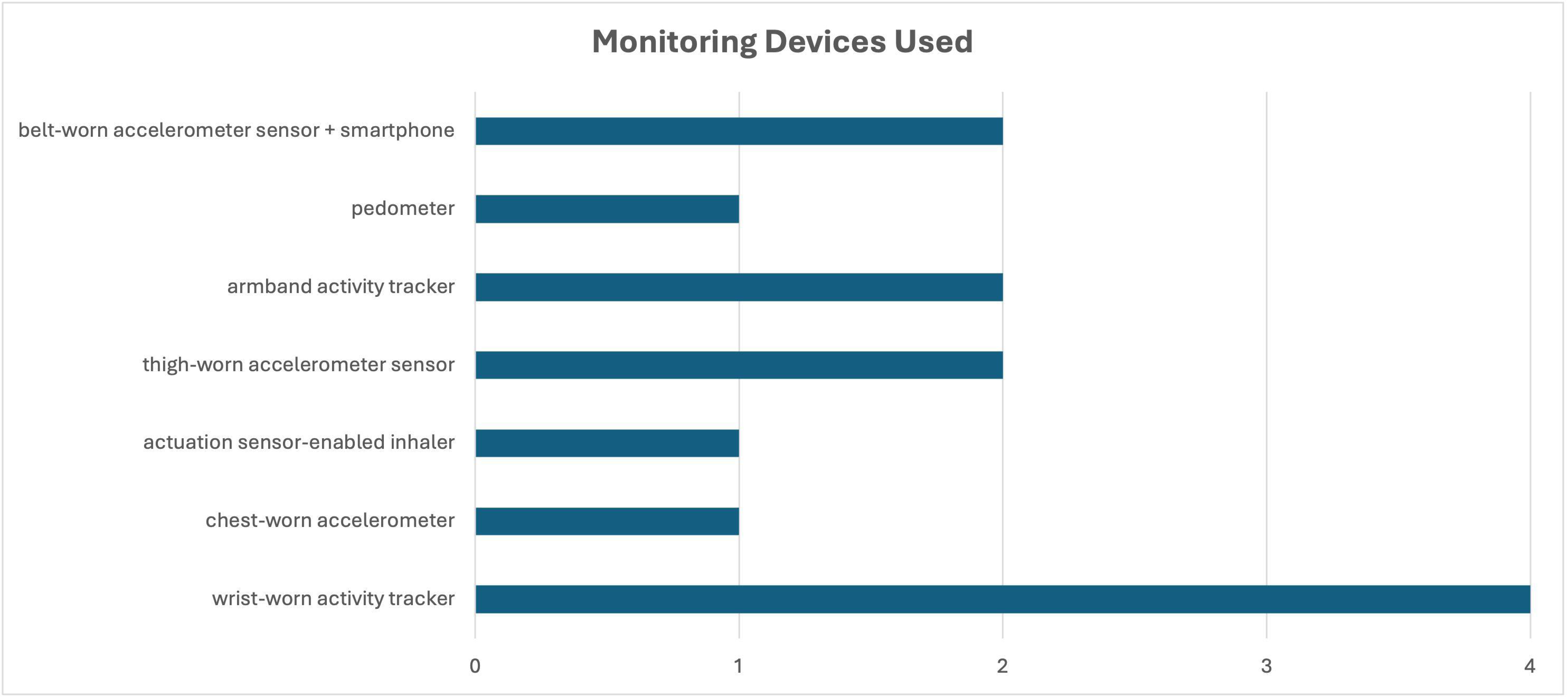
Frequency of Monitoring Devices Used

### What are the most frequent devices used to measure digital endpoints in COPD?

To generate digital endpoints, the most frequently used monitoring strategy employed a wrist-worn activity tracker (n = 4), followed by an armband activity tracker (n = 2), thigh-worn accelerometer sensor (n = 2), and belt-worn accelerometer sensor + smartphone (n = 2). Eight studies used only wearables, and 4 studies used a smartphone plus nearable approach. One study used only a nearable (actuation sensor-enabled inhaler).

### How are digital endpoints used?

Of the 13 studies included, all captured sensor-based data from participants to assess a HBPR program.

Ten studies were RCTs, each with unique HBPR programs [43,45–48,50–54]. Hansen et al (2020) compared the outcomes of two supervised PR programs using a tri-axial accelerometer taped to a participant’s thigh: one at home (n = 67) and another conducted in clinical settings (n = 67) [48]. From baseline to the end of the 22-week follow-up, a statistically significant decrease in steps/day was observed in the conventional PR group (p < 0.05); however, no change was observed in the HBPR group. Using the same accelerometer, Godtfredson et al (2020) examined physical activity level (PAL) differences between HBPR (n = 45) and standard PR (n= 39), using three measures: sedentary time, active time, and steps/day [47]. Twelve months after PR, no significant differences were observed for PAL in the HBPR group but steps/day decreased in the control group (−822 steps/day; 95% CI = [-1575, -138]). Extending these results, Benzo et al (2022) used a wrist-worn activity monitor, demonstrating that a fully remote, unsupervised HBPR program (n = 143) could deliver a significant improvement in steps/day by the end of 12 weeks (p = 0.0113) compared to standard PR (n = 142); however, no clinically meaningful improvements were noted for time spent in moderate-vigorous physical activity (MVPA), light PA, or sleep [43]. Chaplin et al (2022) used a chest-worn accelerometer to compare a conventional PR cohort (n = 51) with a web-enhanced HBPR cohort with enhanced exercise and educational training (n = 52) [45]. While no significant findings were observed in the accelerometer data collected before and after the PR interventions, a nonsignificant increase (12%) in steps/day was observed in the web-based PR cohort (5,645 to 6,112 steps/day), primarily consisting of short MVPA episodes.

One study assessed medication use in a cohort of participants offered HBPR (n = 25), with 75% participating [46]. Federman et al. (2024) used electronic monitoring inhaler devices to quantify medication use remotely in a digital symptom-management platform study, finding that participants in the HBPR program, who were instructed to use their inhaler on a daily basis, exhibited increased daily medication use (difference-in-differences = 7.7%) compared to controls without PR (n = 26) [46].

Two studies used a platform strategy for monitoring, which consisted of a Bluetooth-connected belt-worn accelerometer sensor that paired with a participant’s smartphone, also worn on the belt for the study [51,52]. In the first, Tabak et al. (2014) compared a HBPR program (n = 14) to standard care (n = 16), finding that pedometer-measured steps/day did not improve [52]. In the second, Tabak et al (2014) deployed an autonomous coaching program, that aimed to adjust PA behaviors in real-time via text messaging and visuals, like graphs [51]. Participants (n = 15) responded with appropriate modified behaviors, decreasing PA after a discouraging cue and increasing PA after an encouraging one (p < 0.05). In addition, overall activity levels increased in the cohort; however, although the intervention was designed to more evenly distribute PA across daytime hours, no balance in PA was achieved.

Using a wrist-worn accelerometer, Vasilopoulou et al (2017) investigated the effectiveness of a maintenance HBPR program, one that participants could complete after a primary PR program, if necessary (n = 147) [54]. Considering positive effects with the initial PR program – namely, increased light, lifestyle, and moderate activities and decreased sedentary time (p < 0.05), sustained improvement in all 4 physical activity metrics was observed after the 12-month HBPR program.

Tsai et al’s (2017) study began with an at-home visit to help prepare participants for the HBPR program [53]. This study compared HBPR (n = 19) to a control group (n =17) that received standard care without exercise training, no significant between-group differences in SenseWear armband-measured steps/day were observed at the conclusion of the HBPR. Also using the armband, Horton et al (2021) launched a study with an in-hospital preliminary component to orient participants to the HBPR program [49]. They compared the results of a HBPR program (n = 26) to that of standard, center-based PR (n = 25). Although the HBPR cohort showed promising improvements at the end of 7 weeks, with a significant increase in step count (p = 0.006) and decrease in time spent sedentary compared to the control group (p = 0.039), the step count for both groups reverted to baseline after 6 months. Though no statistically significant change in moderate physical activity (MPA) was observed during the study, time spent in MPA also returned to baseline for both cohorts by the 6-month mark. In contrast, Paneroni et al (2015) used a pedometer to compare the impact of HBPR (n = 18) versus standard PR (n = 18), finding increased physical activity in HBPR participants at study conclusion (3,412 v. 1,863 steps/day; p < 0.001) [50]. Cerdan-de-las-heras et al (2021) applied a wrist-worn accelerometer to track 7-day steps/day and total vector magnitude counts per minute (VMCPM) [44]. No difference from study commencement to end or between HBPR (n = 27) or control cohorts (n = 27) were observed for either digital endpoint.

Benzo et al.’s (2023) small-scale evaluation (n = 12) of a remote coaching program found an overall average increase in monthly step counts, but no statistically significant results derived from monitoring devices; however, individual participant metrics improved over the 8-week study, with 8 participants logging 500 daily step counts [42].

## Discussion

To our knowledge, this is the first scoping review to map digital endpoints used in HBPR for COPD studies. In addition, our objective was to identify the devices most frequently used to capture data for these digital endpoints. Most were published after 2020 and enrolled fewer than 100 COPD patients. While the most common endpoints derived from objective data were daily step count and daily minutes spent in active behavior, multiple studies assessed active behavior by different intensity levels, from lifestyle activities like walking to MVPA. Activity trackers were the most common devices used to capture data, and RCTs were the most commonly used study design. As studies recorded conflicting outcomes, with one showing HBPR participants returning to their lower, baseline PA levels after follow-up periods, it remains to be seen whether self-management programs, implemented after the outpatient PR phase, can sustain the benefits of PR for participants.

Most studies included endpoints capturing physical activity. Given that exercise capacity is an indirect measure of lung function and COPD status, it is not a surprise that most studies evaluated whether an intervention could improve physical activity levels [55]. Increased PA is a key measure for successful completion of PR programs, one that is easy to objectively measure in real-world settings with high-quality consumer wearables [56]. The heterogeneity of study designs and devices used to measure endpoints limits comparative analyses and meta-analyses in this research area; however, if home-based PR programs are ever to be sustainable in the long-run, methodological guidance is necessary regarding the use of wearables.

Across studies comparing steps per day and other physical activity metrics, conflicting results regarding the efficacy of HBPR were found. Given that little is known about participant profiles of those who would benefit most from HBPR, targeted recruitment of studies in COPD monitoring might be necessary to standardize digital endpoints for more efficient interpretation [57]. Though it is important to consider variations in results may be driven by different levels of COPD severity that were not controlled for in analyses, the need for clarity here is critical to inform future studies.

The trend of converging studies around physical activity monitoring is evident. Though valuable for investigating specific targets when evaluating PR programs, this approach disregards the complexity of factors, from environmental to lifestyle behaviors, contributing to home-based PR outcomes, as well as COPD exacerbations and recovery [32,58]. Considering the routine practice of charting lung function at baseline and follow-up for PR program participants in clinical or laboratory environments, it is surprising that no endpoint used a patient-facing spirometer for home-based assessment of lung function. Only the actuation sensor-enabled inhaler was interpreted as an indirect measure of lung capacity, with participants instructed to use their inhaler daily and the endpoint quantifying compliance with daily use guidance. It is possible that some researchers are unaware of modern smart inhaler and spirometer devices or that the reliability of remote spirometer devices has yet to be validated in COPD populations. Regardless, as respiratory disease prevalence grows over the next decade, smart breathalyzers could fill this need in HBPR programs [59].

Similarly, as COPD patients frequently report sleep disturbances, it is remarkable that only 1 study assessed a digital endpoint for sleep (total sleep time) [60]. With an ever-growing number of digital sleep interventions, like digital cognitive behavioral therapy, and digital sleep measures, like nighttime respiratory rate, tracking sleep in COPD programs is ripe for discovery [61]. Also, no digital endpoints evaluated cardiovascular markers, which frequently predict short-and long-term prognosis in COPD [62].

Most studies that met the inclusion criteria paired multiple devices. Despite the variety in devices used, not all data was translated into digital endpoints. For instance, Vasilopoulou et al used a Bluetooth-enabled pulse oximeter, wearable tracker, and novel smartphone app [54]; however, no data from either the pulse oximeter or wearable tracker was converted into a study endpoint. This highlights the gap between available technologies, from Bluetooth-linked inhalers to heart rate-capturing armbands, and clinically meaningful interpretations of the data they generate. Compared to real-world digital endpoints in fields like cardiology, which can involve heart rate and blood pressure endpoints, outcomes evaluated by digital endpoints for HBPR monitoring were primarily restricted to pedometer-derived step counts and accelerometer-derived physical activity [63]. Considering the ever-growing set of digital measures derived from high-quality devices, like nighttime heart rate variability, or emerging digital measures calculated by machine learning techniques, like smartphone audio sensor-derived cough detection, the potential for growth in the home-based PR digital endpoint space is high [64,65].

The results of Benzo et al’s study, which found no significant results between cohorts, but found improved step counts for individuals over their observed time period, support the need for evaluative methods that incorporate patient-specific thresholds in lieu of population reference thresholds [42]. Although using digital endpoints for HBPR monitoring appears feasible, no recommendations regarding optimal tool selection, standardized methods, or sampling frequencies for sensor-based data have been established. Despite daily physical activity being the strongest predictor of survival in COPD patients, the measure is underutilized [66]. The lack of validated or standardized physical activity measures specific to COPD patient populations – and the need for different digital endpoints to help clinicians assess whether individual patients have engaged in distinct types of physical activity, from walking to weightlifting and everything in between, remains a key barrier to evaluating HBPR.

## Limitations

The main limitation is the scope of this review, which necessitated broad inclusion criteria rather than focusing on specific device types or on endpoints. Considering that we limited our study to digital endpoints, this review reflects only the current state of digital endpoints and not the totality of potential endpoints, currently defined as digital measures, that may be in the early stages of development, like audio-based exacerbation measures or EEG headbands to measure sleep [41,65]. Third, our search criteria excluded studies of patients at risk of or with suspected COPD, focusing exclusively on patients with a COPD diagnosis. Thus, other digital endpoints relevant to PR monitoring may exist but may have only been evaluated in the pre-diagnostic phases. We also focused on home-based PR, rather than home-based self-management, and may have excluded digital endpoints that are used in the latter category.

While other digital endpoints relevant to PR monitoring may exist in prediagnostic phases for patients at risk of or with suspected COPD, or for home-based self-management phases, the goal of this study was to focus on HBPR due to the existing gap in knowledge on this phase, as COPD patients in this phase typically being in a higher-acuity state Lastly, we conducted neither a bias nor quality analysis of included articles. Nonetheless, this is the first review study aiming to identify the most common real-world digital endpoints and devices used to evaluate HBPR programs, and to summarize gaps in our understanding of using wearables and nearables in remote monitoring of COPD. Another key limitation inherent in PR programs is that patients with stage 4 COPD are less likely to be referred to PR, whether center-or home-based, potentially restricting the generalizability of these findings to patients with less severe COPD [67]. As the season of monitoring in PR is known to impact participant activity levels but was not reported in studies included in this review, this should be considered when interpreting these results [68].

## Conclusions

Nevertheless, this scoping review revealed that (1) digital endpoints in home-based PR primarily focus on physical activity, (2) digital endpoints tracking sleep, spirometer values, and other cardiovascular biomarkers are understudied, (3) some data streams from wearable devices have yet to be translated into endpoints for home-based PR, and (4) optimal sampling frequency for the digital endpoints in this space is unclear. Future investigations should prioritize robust inclusion criteria for HBPR, to enhance the standardization of digital endpoints by recruiting participants for HBPR who are most likely to benefit, and explore sleep and cardiovascular measures as digital endpoints for home-based PR evaluation [69].

## Data Availability

All data produced in the present study are available upon reasonable request to the authors

## Acknowledgements

Funding for this study was provided by NIH R56 HL173214, NIH R01 HL140486, and the Robert D. and Patricia E. Kern Center for the Science of Health Delivery.

